# Indonesian Version Of The Perceived Stress Scale-10 (IPSS) : A Psychometric Properties Of The Indonesian PSS-10 in Adolescents With Obesity

**DOI:** 10.1101/2025.02.25.25322844

**Authors:** Erlena erlena, Intansari Nurjannah, Deddy Nurwachid Achadiono, Tri Wibawa, Lisa Prihastari

## Abstract

**Introduction:** Stress significantly impacts adolescent obesity, which may impact both physical and mental health outcomes. One popular tool for measuring perceived stress is the Perceived Stress Scale-10 (PSS-10). The purpose of this study is to evaluate the validity and reliability of the Indonesian version of the PSS-10 (IPSS) in this demographic.

**Methods:** Participants of 296 adolescents selected by random cluster sampling, and stratified by geographical location and age. The scale was translated into Indonesian using the translation-back translation technique. For convergent validity, participants filled out the Indonesian PSS-10 and associated psychological tests. The factor structure of the scale was assessed using confirmatory factor analysis (CFA). Cronbach’s alpha was used to determine internal consistency, and a group of individuals was evaluated for test-retest reliability over two weeks.

**Results:** The Indonesian conflict of interest nothing declared PSS-10 demonstrated good internal consistency (Cronbach’s alpha = 0.862) and CFA supported the two-factor model had acceptable fit indices (CFI-0.950, RMR-0.085, RMSE= 0.072) consistent with the original scale, indicating satisfactory construct validity Convergent validity was confirmed through significant correlations with related psychological constructs.

**Discussion:** According to the results, the PSS-10 in Indonesia is a viable and dependable instrument for assessing felt stress in obese teenagers. The tool can be used to understand better the factors associated with stress in this group in both clinical and research settings. Future studies should examine the predictive validity of the scale and how well it applies to other teen demographics.

## Introduction

Adolescent obesity is a growing global health problem. Adolescents who are obese or at risk of obesity face not only physical challenges but also psychological ones, including increased stress levels. High stress in obese adolescents can affect eating behaviour, such as increased fat intake, which further worsens obesity(1). A variety of factors, such as academic proficiency, social changes, and technological advancements, contribute to the rise in stress levels in the adolescent community. According to research conducted in Indonesia, the prevalence of general mental disorders (CMD) is very high among the elderly, with the prevalence being higher among the elderly than among children(2). n addition, research indicates that obese people are more likely to sleep experience, fatigue, and an increase in cortisol levels, which are physical indicators of chronic stress (3).

Valid and reliable instruments are needed to lower the stress level in adolescents. One tool that is frequently used is the Depression Anxiety Stress Scale (DASS). However, the validity and reliability of this tool must be carefully examined to ensure that it is in line with the Indonesian populace (4). According to a study, there is a lack of adequate validation among Asian populations on the use of DASS-21 in Asian samples, including Indonesia. This kind of cultural variation might affect people’s emotional expression and personal experiences (5). Alongside this, adjustments and validations have been made to online learning environments during a pandemic, such as the Student-Life Stress Inventory (SLSI). The COVID-19 virus (6).

This study highlights how important it is to adjust the measure to the current conditions and context that are experienced by adolescents. The development and validation of stress measurement instruments that are appropriate to the conditions of Indonesian adolescents is important to ensure the accuracy and relevance of measurement results. This will help in designing appropriate interventions to address stress issues in adolescents. The Perceived Stress Scale (PSS), developed by Cohen, Kamarck, and Mermelstein in 1983 (7), is a widely used tool to measure a person’s stress level. The PSS-10, a shortened version of the PSS consisting of ten items, is a valid and reliable tool for assessing a person’s perception of stress under general conditions during the past month (8). To ensure that the tool can be used appropriately across different groups of people, it has been translated and validated in multiple languages (9). the cultural, linguistic, and socio-economic backgrounds of Indonesian society are very diverse. Therefore, to ensure their validity and relevance, stress measurement instruments such as the PSS-10 must be adapted to the Indonesian cultural and linguistic context. For obese adolescents, the use of a valid Indonesian version of the PSS-10 can provide in-depth insights into how stress affects their well-being. Validation of the PSS-10 for obese adolescents is an important step in supporting stress measurement and can be used to build multidisciplinary intervention programs that include physical, psychological, and social elements, and encourage more research focused on the relationship between stress, eating behaviours, and obesity in this age group (10). Reliable measurement tools are needed to design more effective interventions and policies to improve the physical and mental health of obese adolescents (11).

Several studies have used the Perceived Stress Scale (PSS-10) to examine the relationship between stress, obesity, and other psychosocial factors. This study examined the factor structure and validity of the PSS-10 in 1,574 Chinese adolescents(12). Research has indicated that perceived stress is associated with emotional eating in adolescents. A study titled “Perceived Stress and Emotional Eating in Adolescence: Mediation Through Negative-Focused Cognitive Emotion Regulation and Reward Sensitivity” found that higher perceived stress levels were linked to increased emotional eating behaviours, which can contribute to obesity risk (13). These studies highlight the importance of addressing psychosocial factors, including perceived stress and social pressures, in interventions aimed at reducing obesity risk among adolescents (14).

PSS has been translated into many languages, such as Indian (15), Persian (Iran) (16), Chinese (17), Germany (18), Philippines(19) even though many researchers have verified the Perceived Stress Scale in various languages, there is not enough evidence for the Indonesian version. The purpose of this study was to evaluate the psychometric aspects of the Indonesian Perceived Stress Scale (IPSS). This study focusing on the psychometric validation of the PSS in obese or at-risk adolescents is urgently needed. This will ensure that the measuring instrument is effective in identifying stress levels so that appropriate interventions can be implemented to improve adolescents’ mental and physical health.

## Method

### Participant

Participants in this test were early adolescents aged male and female 13 to 15 years. research design using cross sectional study. Participants were recruited by random sampling for 4 school, each student filled out a stress level questionnaire accompanied by the researcher, the recruitment period began for 3 weeks from October 02, 2024, to October 22, 2024. participants provided informed consent by each parent in writing and is collected and explained at one time at each school.To see demographic data See table 1

**Table 1.**
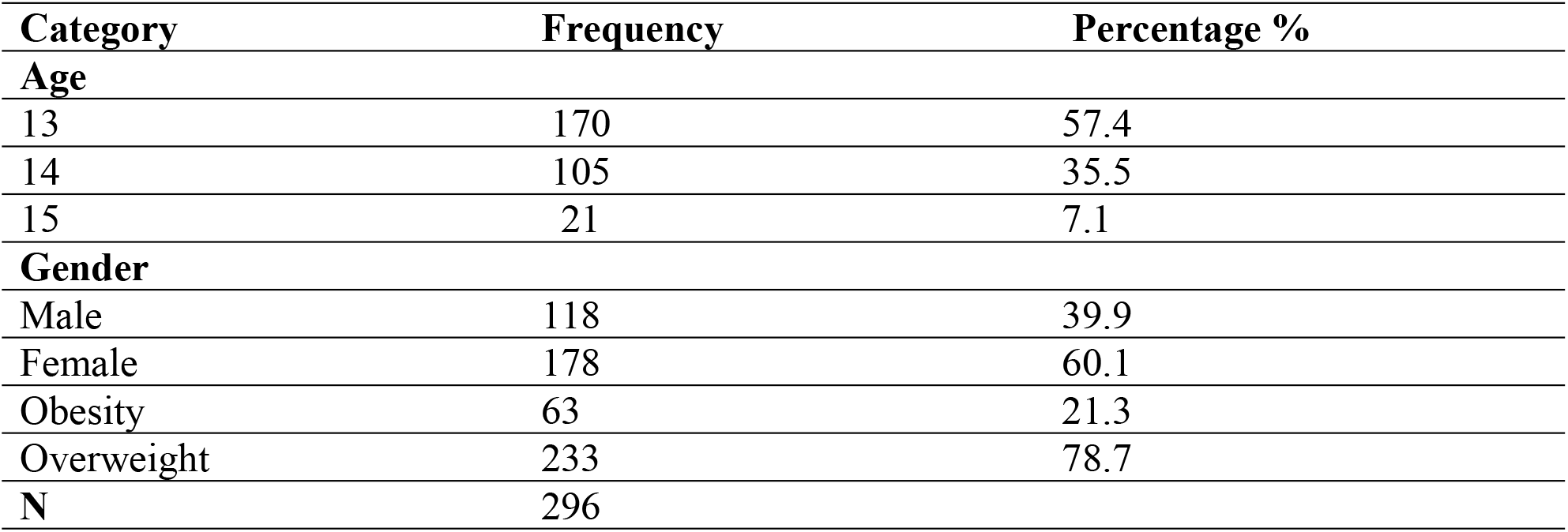
Demographic data.

| Category | Frequency | Percentage % |
| --- | --- | --- |
| **Age** |  |  |
| 13 | 170 | 57.4 |
| 14 | 105 | 35.5 |
| 15 | 21 | 7.1 |
| **Gender** |  |  |
| Male | 118 | 39.9 |
| Female | 178 | 60.1 |
| Obesity | 63 | 21.3 |
| Overweight | 233 | 78.7 |
| **N** | 296 |  |

### Translation Process

The PSS-10 was modified externally and back-translated by adding four English language experts and three adolescent psychologists who took the readability test. The translation of the scale began with a forward translation from English to Indonesian by two different translators before synthesis. Then, two different translators translated the synthesized results back into English(20). (stage 1),

In the next step, a team of experts consisting of researchers and psychology professors studied the concept of stress (21) (stage 2), see table 2. After considering suggestions and corrections, the inventory of Indonesian language forms is ready for initial testing. Thirty people were tested on readability. The purpose of the readability test was to provide information about words in sentences that were difficult for readers to understand (22). (stage 3), see fig 1

**Table 2.** The content Validity Indexes.

| ITEM | Expert 1 | Expert 2 | Expert 3 | Expert in<br>Agreemen<br>t | I-CVI | UA |
| --- | --- | --- | --- | --- | --- | --- |
| 1 | 1 | 1 | 1 | 3 | 1,00 | 1 |
| 2 | 1 | 1 | 0 | 2 | 0,67 | 0 |
| 3 | 1 | 1 | 1 | 3 | 1,00 | 1 |
| 4 | 1 | 1 | 1 | 3 | 1,00 | 1 |
| 5 | 1 | 1 | 1 | 3 | 1,00 | 1 |
| 6 | 1 | 1 | 1 | 3 | 1,00 | 1 |
| 7 | 1 | 1 | 1 | 3 | 1,00 | 1 |
| 8 | 1 | 1 | 1 | 3 | 1,00 | 1 |
| 9 | 1 | 1 | 1 | 3 | 1,00 | 1 |
| 10 | 1 | 1 | 1 | 3 | 1,00 | 1 |
|  | 1 | 1 | 0,9 | S-CVI<br>/Ave | 0,97 |  |
| Proportion<br>relevance | 1,1 | 1,1 | 0,99 | S-CVI/UA |  | 0,9 |
| The average proportion of items judged as relevant across the<br>three experts |  |  |  |  | 1,1 |  |

**Fig 1.**
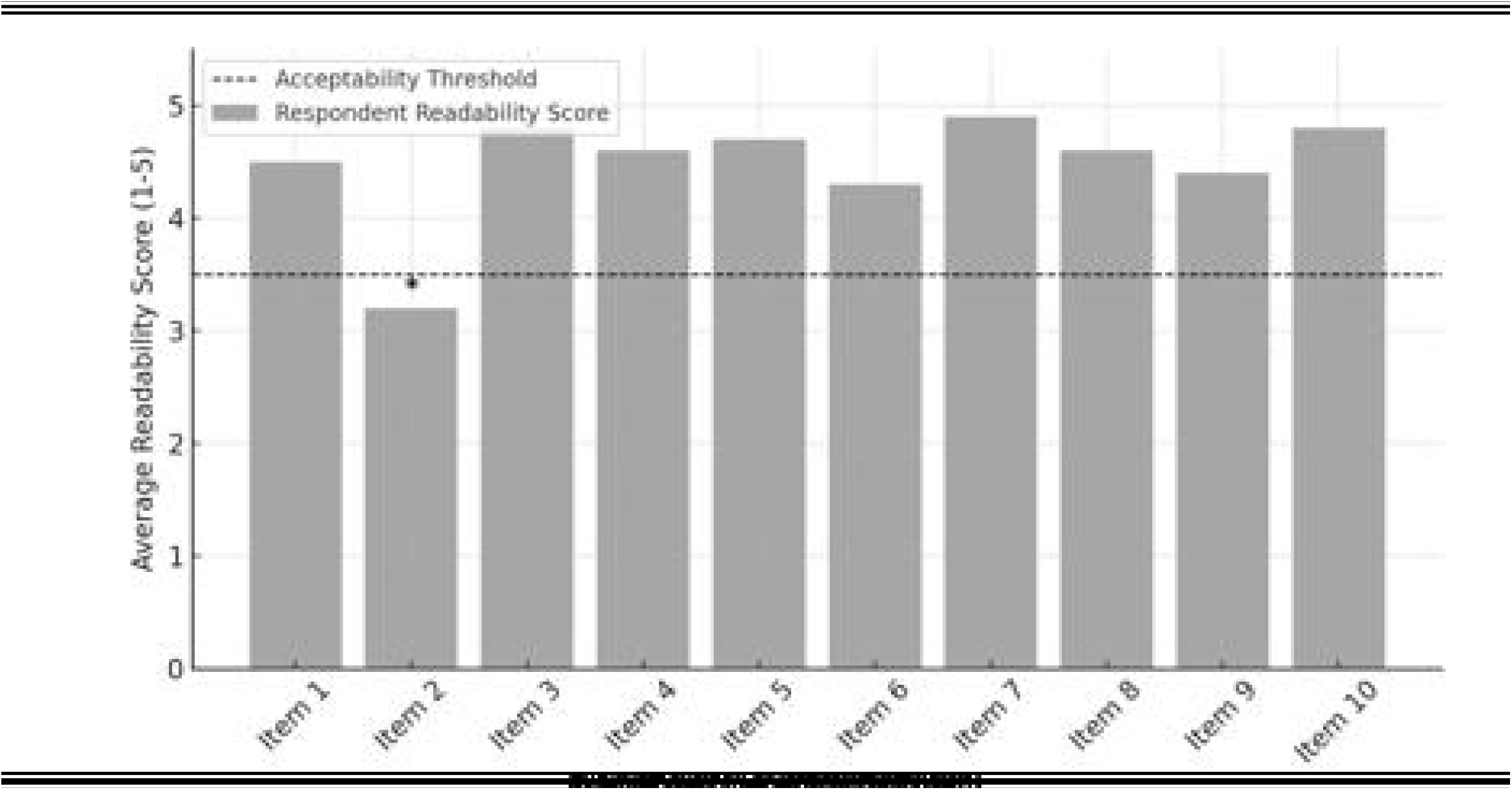
Readability Scores.

In the next stage, the researcher conducted a validity and reliability test on 30 adolescent respondents to assess the level of validity and reliability of this questionnaire (23). (stage 4)

In this study, the Perceived Stress Scale (PSS-10), created by Cohen et al., consists of two main dimensions: perceived incompetence and perceived ability. In this study, there are ten items rated on a five-point Likert scale, where 0 indicates never, 1 indicates rarely, 2 indicates sometimes, 3 indicates quite often, and 4 indicates very often(24).

This study was approved by The Medical and Health Research Ethics Committee (MHREC) Ref No; KE/FK/1097/EC/2024, The entire study was carried out by the principles of the Helsinki Declaration (version 2013).

### Data collection

Data for this study were collected through a scale administered to adolescents. The study involved 296 adolescents, and the sample size met the criteria outlined by Kennedy (25), which requires a minimum of 100 people(26). The sample size adequacy criteria are as follows: 50 is very inadequate; 100 is inadequate; 200 is adequate; 300 is sufficient; 500 is very sufficient; and 1000 or more is very satisfactory(27). This study used continuous sampling, with questionnaires given to adolescents who were willing to complete the scale. Data were collected over a 3-weeks adolescents were eligible for inclusion (criteria included written informed consent, not participating in another survey, Adolescents were selected by random cluster sampling, and stratified by geographical location and age. The sample size for the IPSS was calculated with a confidence level of 95% and ± 0.3 error, based on screening the variance in body mass index, and age. A sample size 296 for 4 school.

### Statistical Analysis

Data were analyzation SPSS® (version 22) and SEM™ statistical software. Descriptive statistics were applied to analyses demographic information and features. The factor structure was examined using Confirmatory Factor Analysis (CFA), which is a suitable method for testing theory-driven models and validating the structure of psychometric instruments(28),(29). See table 3

**Table 3.** Loading Factor in the CFA Model of PSS-10.

|  |  | Estimate | SE | Value-<br>p | Initial<br>Eigenvalue | Variants<br>%<br>Explained | Cumulative<br>% Variance | Coefficient<br>y Cronbach |
| --- | --- | --- | --- | --- | --- | --- | --- | --- |
| Factor 1 | Item2 | 1.000 |  | < .001 | 4,495 | 44.95 | 44.95 | 0,876 |
|  | Item1 | .879 | .065 | < .001 |  |  |  |  |
|  | Item3 | .949 | .074 | < .001 |  |  |  |  |
|  | Item10 | 1.013 | .083 | < .001 |  |  |  |  |
|  | Item9 | 1.021 |  | < .001 |  |  |  |  |
| Factor 2 | Item5 | 1.000 | .065 | < .001 | 2.138 | 21.38 | 66.33 | 0,857 |
|  | Item4 | .962 | .061 | < .001 |  |  |  |  |
|  | Item6 | .853 | .066 | < .001 |  |  |  |  |
|  | Item7 | .868 | .068 | < .001 |  |  |  |  |
|  | Item8 | .861 | .079 | < .001 |  |  |  |  |

The Kaiser-Meyer-Olkin (KMO) measure and Bartlett’s Test of Sphericity were used to assess the suitability of the dataset for factor analysis. Model fit was evaluated using multiple indices, including the chi-square value (with a significance level of α = 0.05), degrees of freedom, Comparative Fit Index (CFI), Standardized Root Mean Residual (SRMR), and Root Mean Square Error of Approximation (RMSEA)(30). The Content Validity Index for items (I-CVI) and the scale (S-CVI) were calculated by those who define content validity as the extent to which an instrument’s items are representative of the construct being measured The I-CVI was computed by dividing the number of experts who agreed on the relevance of an item by the total number of experts, while the S-CVI was obtained by calculating the mean of all I-CVI values. The Pearson Correlation Test was performed to evaluate the relationship between factors and to establish criterion-related validity(31). See table4

The majority of participants were **13-year-old adolescents (57.4%)**, with a higher proportion of **females (60.1%)** than males.

Table 1 represents the content validity indexes calculated after the expert ratings. In line with our results, Beaton et al. (7) suggested that the responses and answers to the interview ensured that the adapted version maintained the equivalences. Participants rated how well they understood each item by scoring between 0 and 1 (agrees and disagrees). in this study, experts were asked to rate the relevance of each item to IPSS. as it was advised (10, 32), three experts rated the items between 0 and 1, the I-cvi (≥0.67 for all items, with 90% achieving 1.00) suggests that the majority of items received full agreement from experts, reinforcing their relevance in measuring Indonesia perceived stress scale, the s-cvi/Ave (0.97) and S-cvi/Au (0.90) exceed the recommended 0.80 threshold, meaning that the overall scale has strong content validity, making it suitable for assessing stress perception. Based on the expert ratings, item 2 received a score of 0 from one expert, while the other two experts rated it as 1. This resulted in an item-level content validity index (I-CVI)of 0.67, which is below the commonly accepted threshold of 0.78 for three experts. Since the agreement among experts on this item is not strong, the researcher re-evaluated and revised item 2 to improve its clarity, relevance, or alignment with the construct being measured. The researcher reviewed expert feedback, rewriting the item, adjusting the wording to make it clearer and more precise. Reassessing the content fit. Ensuring that the item aligns with the overall construct and purpose of the scale. Revalidating the item and conducting another round of expert review to confirm improvements.

### Factor Loading (Estimates)

The two-dimensional construct was confirmed by conducting a CFA approach to ensure whether the construct indicators are valid indicators of the latent construct. The analysis results of all indicators in Factor 1 and Factor 2 have significant factor loadings (p < 0.001). Factor 1: Item 2, Item 1, Item 3, Item 10, and Item 9 have factor loadings ranging from 0.879 to 1.021. Factor 2: Item 5, Item 4, Item 6, Item 7, and Item 8 have factor loadings between 0.853 and 0.962. See fig 2. This suggests that all items have strong Loading Factor values (>0.5) and can be retained, Standard Error (SE) shows the variability of parameter estimates, with fairly small values indicating stability in the estimates, Variants explained by Factor 1 contribute 44.95% of the total variance, making it the most dominant, factor 2 contributes 21.38% of the total variance, which is still significant. Together, these two factors explain 66.33% of the total variance, meaning they capture most of the data’s structure. Coefficient Cronbach’s Alpha factor 1 and factor 2: 0.876 and 0.857 shows high reliability, which means the internal consistency of the items in the factor is quite good.

**Figure 2.**
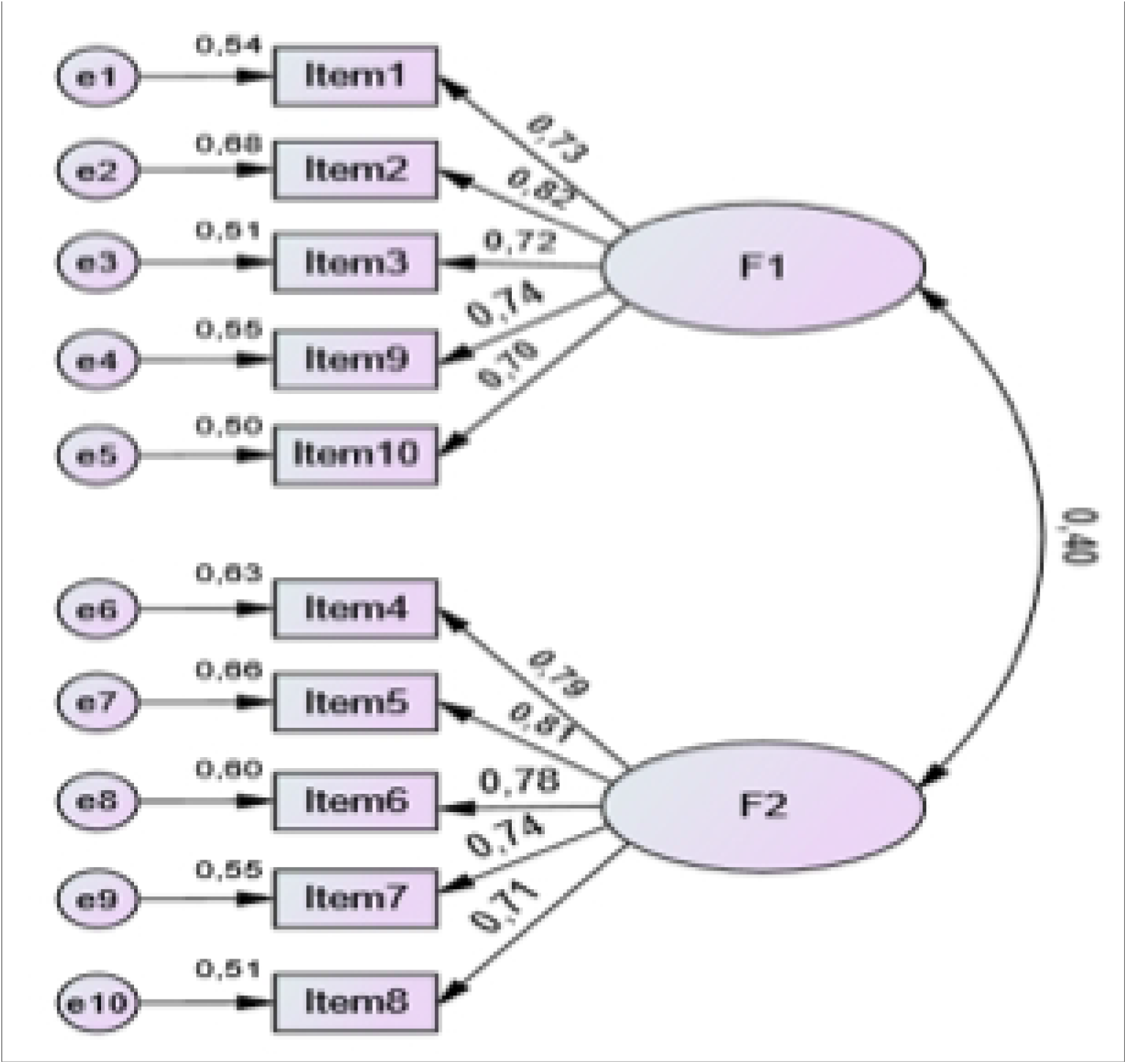
Two-factor IPSS-10 CFA Model.

**Table 4.** Goodness-of-fit indices of two CFA models of the IPSS-10 (n = 296)

| Fit Indicator | Criteria | Research Result | Information |
| --- | --- | --- | --- |
| NFI | >0,940 | >0,90 | Fit |
| RMSEA | <0,08 | 0,072 | Fit |
| CFI | >0,90 | 0,963 | Fit |
| TLI | >0,90 | 0,950 | Fit |
| RMR | <0,08 | 0,085 | Almost good, slightly above the threshold |

The Goodness of Fit (GOF) results for all aspects meet the criteria. For CFI >0.9, The model fits the data well, as indicated by multiple goodness-of-fit measures (NFI, CFI, TLI, RMR), The RMSEA value (0.072) is within an acceptable range (**< 0.08**), indicating a good fit based on indicator, but it is near the upper threshold, suggesting some room for improvement.

## Data Availability

All relevant data are within the manuscript and its Supporting Information files.

## DISCUSSION & CONCLUSION

The analysis conducted evaluates the readability, validity, and reliability of the measurement instrument used to assess stress in adolescents. The results from statistical analysis, including factor analysis and expert validity assessment, provide insight into the robustness of the instrument.

The PSS scale measures how individuals perceive stress and their ability to cope. The two identified factors align with previous research, where stress instruments often distinguish between Perceived distress (Factor 1): Feelings of being overwhelmed, helpless, or out of control. Coping ability (Factor 2): The extent to which an individual feels they can handle stress effectively. Given that the respondents are adolescents, developmental psychology theories (e.g., Erikson’s psychosocial development) suggest that this age group is particularly vulnerable to academic pressure, social relationships, and identity formation stress. Therefore, ensuring that the instrument accurately captures these experiences is critic. Factor 1 accounts for 44.95% of the variance, while Factor 2 accounts for 21.38% of the variance, leading to a cumulative explained variance of 66.33%. According to Hair et al. (2010), a cumulative variance above 60% is generally considered acceptable in psychological and social science research, indicating that the identified factors adequately capture the construct being measured. Factor loadings reveal that items 1, 2, 3, 9, and 10 strongly load onto Factor 1, while items 4, 5, 6, 7, and 8 load onto Factor 2. This suggests that the questionnaire likely assesses two distinct dimensions of stress in adolescents, potentially perceived stress and coping strategies, as commonly identified in psychological stress measurement tools such as the Perceived Stress Scale (PSS) developed by Cohen et al. (1983). The Cronbach’s alpha for Factor 1 (0.876) and Factor 2 (0.857) suggest good internal consistency, as values above 0.70 are considered acceptable (Nunnally & Bernstein, 1994). This indicates that the items within each factor are measuring a cohesive underlying construct, which supports the reliability of the instrument. The measurement instrument demonstrates strong validity and reliability, making it suitable for assessing adolescent stress. However, Item 2 requires revision to improve content validity. The findings align with stress measurement theories, reinforcing the importance of psychological tools tailored to adolescent experiences. Future research should focus on refining the scale and ensuring it remains relevant across diverse adolescent populations

## Acknowledgment

The authors would like to acknowledge the Faculty of Medicine Gadjah Mada University Indonesia Endowment Funds for Education (LPDP) Centre for Higher Education Funding (BPPT)

## Author Contributions

Conceptualization, Writing, Data curation Ee, IN

Formal Analysis, LP

Investigation IN

Methodology Ee

Project administration TW

resources, Ee,LP

Validation DN

Visualization DN

## Suporting Information

S1 Fig. Fig 1. Readability Scores

S1 Table. Demographic data

S2 Table. The content Validity Indexes

S3 Table. Loading Factor in the CFA Model of PSS-10

S4 Table. Goodness-of-fit indices of two CFA models of the IPSS-10

